# Stigma among people living with HIV in Northern Egypt

**DOI:** 10.1101/2024.08.27.24312679

**Authors:** Mona Magdy, Adel Zaki, Sherif Omar Osman, Ekram W. Abd El-Wahab, Asmaa Abd Elhameed

## Abstract

Stigma involves labeling individuals, and associating them with negative behavior, leading to social isolation, stereotypes, loss of status, and discrimination. HIV stigma is considered a major barrier to effective responses to the HIV/AIDS epidemic. We aimed in this study to assess the stigma among people living with HIV (PLWH) in Alexandria, Egypt. We conducted a cross-section study on 785 PLWH attending the HIV unit in Alexandria Fever Hospital. Data collection was done through an interviewing questionnaire and an Arabic-translated version of the standardized, 12-item short version of the HIV Stigma Scale. The stigma scale was highly reliable (alpha coefficient = 0.743). About 72.4% of participants had high stigma with a total stigma score of more than 30. Men showed a higher median stigma score than women (60.8% vs 23.0% respectively recorded a median score of 33 or more). The total stigma score was significantly associated with the male sex and residing in Alexandria city (p= <0.1).

In conclusion, PLWH in Egypt experience a significant degree of stigma. These data can guide future health interventions to provide psychological and social support for PLWH.

## Introduction

The concept of stigma is not of recent origin. At its essence, stigma can be described as a “mark” or an attribute of oneself that society devalues [1] and it encompasses the act of labeling individuals or groups and linking these labels to undesirable actions. Individuals who are labeled in this way stand out from the broader societal norm, encounter stereotypes, undergo a loss of social standing, and are subjected to discriminatory practices [2].

HIV-related stigma manifests in diverse manners, encompassing anticipatory stigma, internalized stigma, and enacted stigma [3,4,5] which all result in silence and denial, self-blame, rejection, violence, self-isolation, failure to disclose one’s HIV status, and secondary stigma of HIV services providers [6]. As a result, accessing or consuming HIV medications can produce anxiety over potentially revealing one’s HIV status to others and heightens mindfulness of one’s disease [5]. A growing body of evidence points to a link between HIV stigma and its unfavorable impact on individual’s quality of life and adherence to anti-retroviral therapy (ART) [7,8].

HIV landscape in the MENA is shaped by diverse socio-economic and political factors which have posed substantial challenges in addressing public health issues, including HIV [9,10]. Egypt is categorized as having the lowest HIV prevalence rates in the region, as measuring less than 0.1% in 2020 [11] with an annual increase in people living with HIV (PLWH) by 20 to 25% [12].and an estimated ART coverage of 40% [37 - 45%] [13].

Addressing stigma to HIV/AIDs in healthcare settings in Egypt as a priority to end the HIV epidemic by 2030 [14]. The demographic health survey conducted in Egypt in 2014, revealed a universal attitude of stigma and discrimination towards HIV/AIDS within Egyptian society [15]. Updated data on stigma among PLWH is critically needed to evaluate a stigma-reduction and guide evidence-based operational interventions and policy making.

## Methods

### Study design, setting and population

We conducted a cross-sectional study between January to December 2021 at the main Fever Hospital in Alexandria governorate, a city in north of Egypt. Alexandria Fever Hospital is a pooling hospital that serves inhabitants from Alexandria and nearby governorates. The target population was PLWHA attending the HIV clinic at Alexandria Fever Hospital as a centralized center for dispensing ART. Potentially eligible participants were defined by as adults aged 18 years or older, mentally well, on ART as of 31 December 2020, and voluntarily willing to participate and sign an informed written consent. Those who were mentally ill, incarcerated (being inaccessible to the researcher), and HIV-positive cases unaware of their health status (yet undisclosed by the health care team) were excluded from the study.

The sample size was calculated by the PASS 12 program[16] based on a level of adherence to ART among PLWHA (86.0%) revealed in a pilot study conducted by the researchers on 40 subjects that were not included in the final analysis. Using a 95% confidence interval (C.I.), a degree of precision equal to 2% and a level of significance set at 0.05, the calculated sample size was equal to 559, however, 785 adult PLWHA were eventually enrolled in the study.

All PLWH attending the HIV clinic at Alexandria Fever Hospital were invited to participate in the study. those who accepted to participate and signed an informed written consent got a more detailed description of the study aim and procedure.

We excluded from the study PLWH who were mentally ill, incarcerated, those who were yet unaware of their HIV seropositive status.

### Data collection

A face-to-face interview questionnaire was designed as the primary data collection instrument, initially constructed in Egyptian Arabic and administered in a printed form and distributed personally to the participants by the researcher. Each interview took around 10-15 minutes to complete. The questionnaire consisted mostly of closed-ended questions and a few open-ended inquiries, to provide more diverse details.

The first part of the questionnaire served to collect data about sociodemographic characteristics (age, sex, marital status, residence, education, employment, and HIV risk behavior). The second part of the questionnaire included questions of the HIV Stigma Scale.

### Stigma measurement tool

HIV stigma is highly related to society, and it is culture. Thus, the content and expression of stigma vary across the globe. Therefore, tools to measure HIV stigma should be culturally sensitive. The 40-item “HIV Stigma Scale” is one of only a few instruments that covers all stigma processes affecting individuals with HIV and is the most widely used of the various tools meant to measure HIV stigma [17]. So, a shorter easy to administrate instrument was created to assess HIV stigma in larger surveys. We used the short version of the HIV Stigma Scale developed by Reinius et al [18], which was proved to be valid, and has comparable psychometric properties to the full-length scale.

Stigma scale questions (numbers 14 – 25) are divided in 4 subscales (Personal stigma, Disclosure concerns, Concerns about public attitudes, and Negative self-image). For each question a 4-point Likert scale was set to include a score range from 4 “always”, 3 “sometimes”, 2 “rare”, and 1”never happened”. Responses were summed to calculate the possible final score for each subscale with a possible total score range from 3 to 12. Higher scores reflect a higher level of perceived HIV-related stigma.

### Translation of the 12-item short version of the HIV stigma scale

The original version of the Stigma scale was available and free to use without prior permission. The Arabic version is the same structure and sequence as the original English version. The WHO Guidelines were followed in the translation. The translation steps included (Forward translation, Reconciliation, Back-translation, Pre-testing and cognitive interviewing and Final version)[19].

### Internal consistency of the short version stigma scale

The internal consistency (reliability) of the translated short version stigma scale was measured by coefficient alpha (α).

### Pilot study

A pilot study was conducted on 40 PLWH (10% of the intended sample) to assess the feasibility of conducting the study, test the data collection tool, and identify any questionnaire design issues. Cloudy items were identified, necessitating modification. All data from the pilot was later omitted from the final study to ensure both validity and reliability.

The pilot test revealed the cloudiness of some items. For example, the stigma scale question number 17: “I am very careful who to tell that I have HIV?”, the statement “at work” was added at the end of the question, to measure how the work community would accept a co-worker living with HIV. Nevertheless, in question number 21: “Do most people believe that a person who has HIV is dirty?”, the word “dirty” was replaced with “badly behaved” to make it more acceptable in our culture.

### Statistical Analysis

Data were structured and processed with SPSS version 22. Descriptive statistics, including frequencies and percentages were used for categorical variables. Medians and interquartile ranges (IQRs) were used to present continuous variables. Bivariate analyses (Mann-Whitney, Kruskal Wallis test) were run to examine the association between the total stigma score and independent variables were computed and presented.

The reliability of the tool used to assess the internal consistency of the stigma scale was tested using Cronbach alpha. In all statistical tests p values <0.1 was set as a level of statistical significance.

## Results

### Sociodemographics of the enrolled PLWH

The study included 785 participants. The majority were male (72%), in the age group 30-49 (67.8%) with a mean age of 37.37 ± 10 years, residing in Alexandria (85.1%), educated (88.5%), and having fulltime employments (52.2 %). The reported modes of acquiring HIV were more through heterosexual transmission (having multiple sex partners) (53.1%), intravenous drug use (IVDU) (13.5%), engagement in homosexuality (12.7%), and blood transfusion (6.2%) (Table 1).

### Description of the 12-short version of the HIV stigma scale among the enrolled PLWH

The “personal stigma”, the “concerns about public attitudes”, the “disclosure concerns” and the “negative self-image” subscales showed median scores of 3 (IQR= 3–5), 12 (IQ=12–12), 12 (IQ= 8-12) and 9 (IQR= 7–11) respectively. The “personal stigma” subscale was the most impacted as 370 (47.1%) of the participants did not answer the whole subscale because they did not tell people around them about their disease (Table 2 and Figure 1).

**Figure 1.**
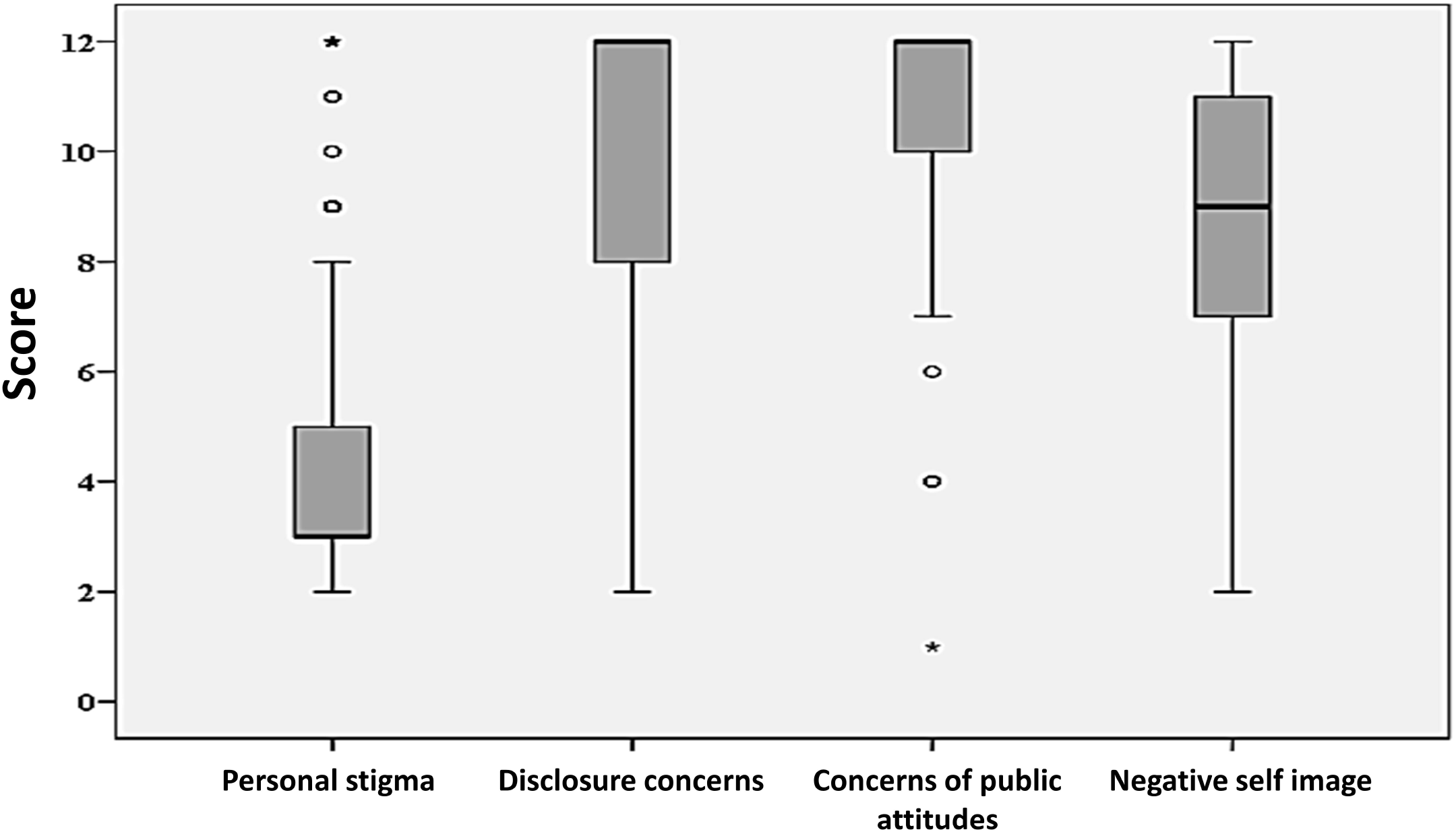

As a measurement of the internal consistency, the short version stigma scale had an alpha coefficient (α) of 0.743 (Table 2), indicating good reliability of the scale. However, the internal consistencies of individual subscales were higher [“personal stigma”; α = 0.94, “disclosure concern”; α = 0.88, and “concerns with public attitudes”; α = 0.87], with the exception of the subscale “negative self-image” where α = 0.657.

The total stigma score was significantly associated with the male sex and residing in Alexandria city (*p*= <0.1) (Table 3). Men revealed a higher median score than women in the “Disclosure concerns” and “Negative self-image” subscales, while both men and women showed the highest median score of 12 in “concerns about public attitudes” subscale, and equally had a low median score of 3 in the “Personal stigma” subscale (Figure 2). Regarding the total median stigma score, 60.8% of men and utmost 23.0% of women had a high median score of 33 or more. A high median score of 12 in both the “disclosure concerns” and “concerns about public attitudes” subscales was found in the age group 30–49, while a median score of 12 in the “concerns about public attitudes” subscale was more achieved by those above 50 years of age (Figure 3).

**Figure 2.**
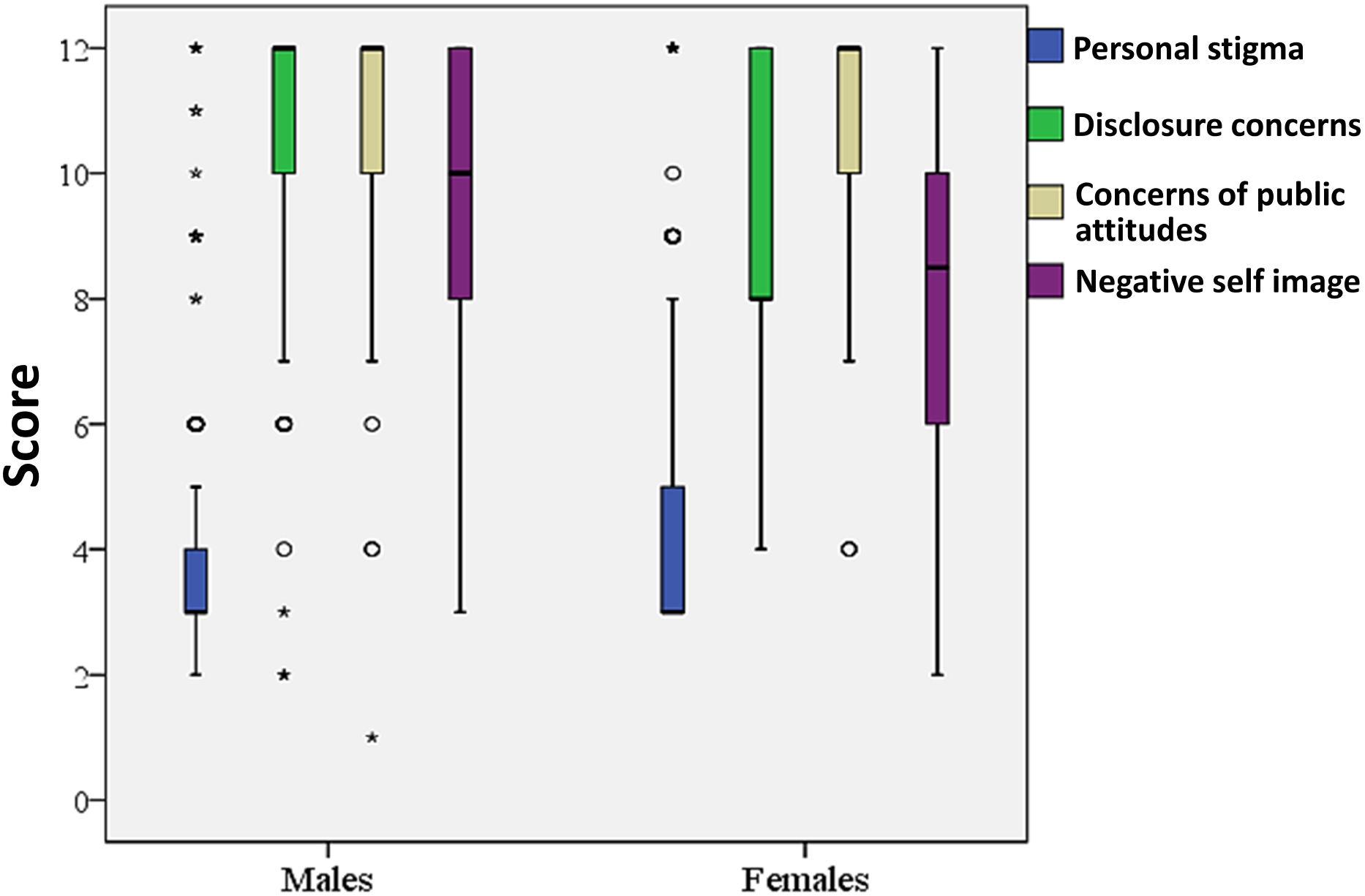

**Figure 3.**
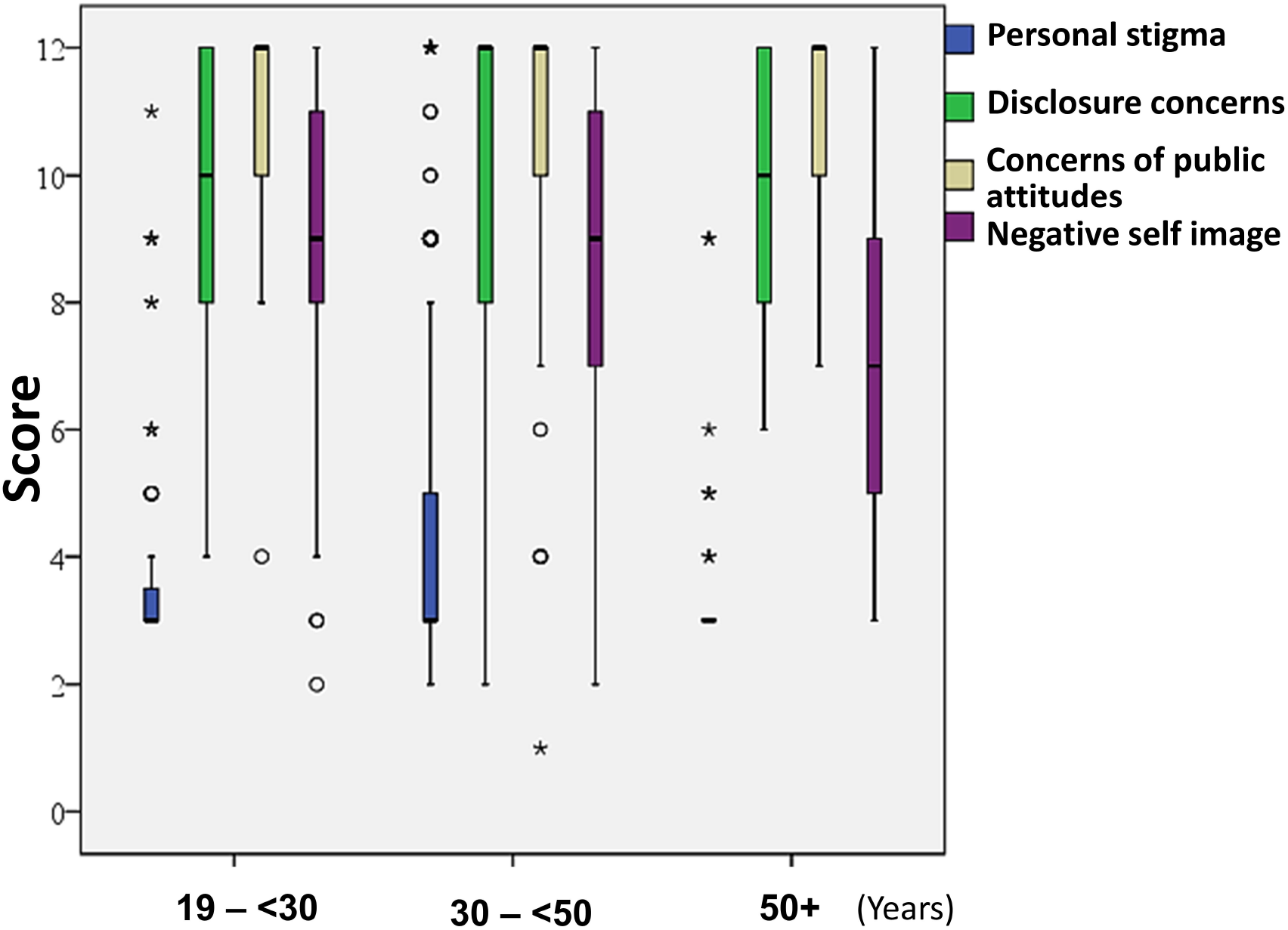

In the “personal stigma” subscale, the highest scores (i.e., represented high level of stigma) were responses to questions 14 and 15, stating: “Some people avoid touching me once they know I have HIV”, and “People I care about stop calling after knowing that I have HIV”, as 3.1% of the responders answered, “always”. While in the “disclosure concerns” subscale, the highest score was in question 19 “I am very careful about telling anyone I have HIV (work)”, as 97% of the responders answered “always”. In “concerns about public attitudes”, the highest score was for question 21 “Most people believe the person with HIV is a bad person”, as 92.6% of the responders answered “always”. Finally, in the last subscale

“negative self-image” the highest score was in question 24 “People’s attitudes toward HIV make me feel worse about myself”, 67.1% of the responders answered “always” (Figure 4).

**Figure 4.**
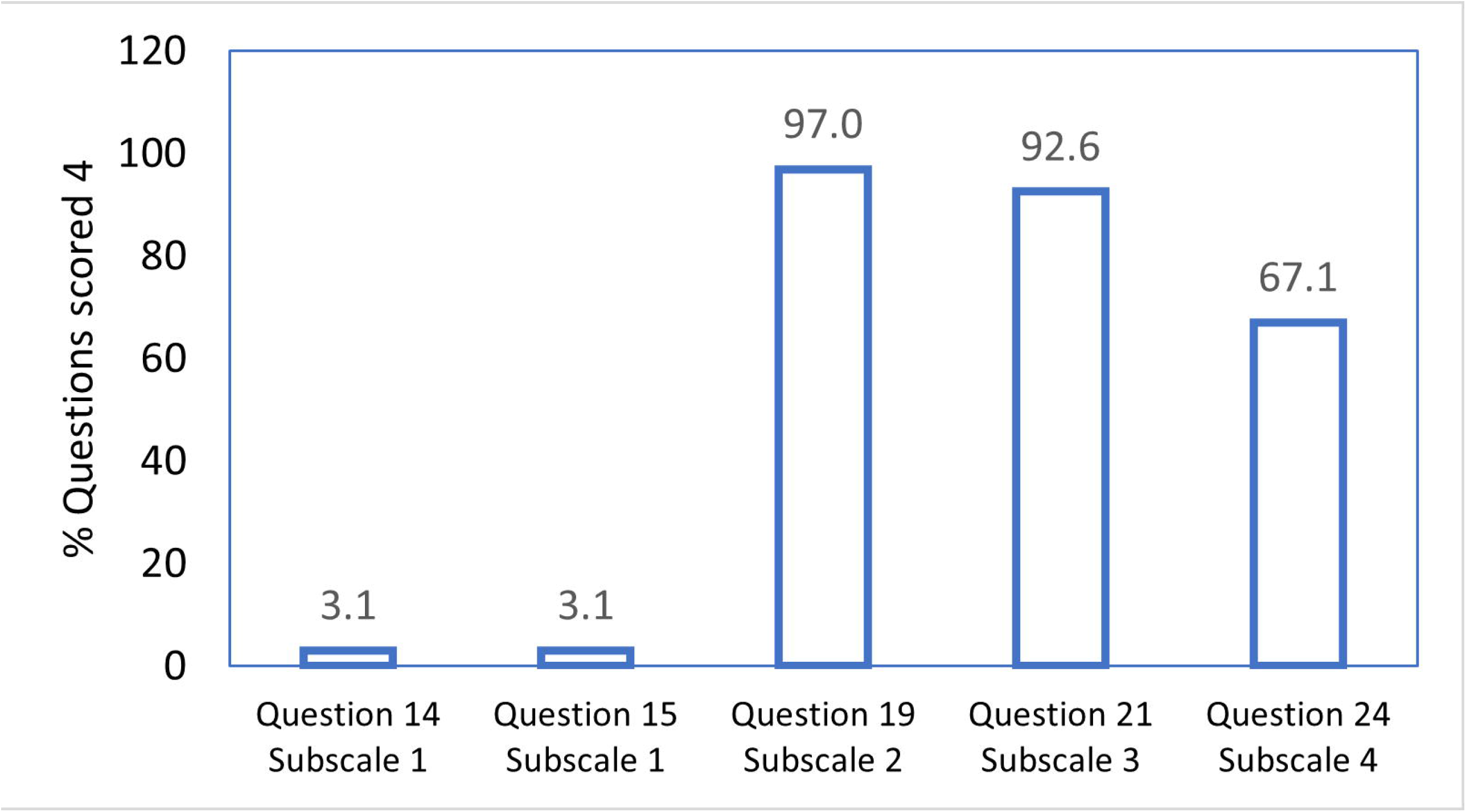

## Discussion

The present work revealed a high stigma score (more than 30 out of 48 the total stigma score), among PLWH who participated in the study. A significant number of the participants did not disclose their illness to their relatives and friends due to their concern about the reactions of others and the potential for stigma and discrimination.

Many studies revealed the importance of disclosing HIV-positive status as it plays a crucial in social support and enhancing adherence to ART [20,21,22,23,24]. Certainly, PLWH who have revealed their status to their families are able to take their HIV medications fearlessly [25,26].

The majority of the PLWH participants in the present study showed a higher score in “ Negative self-image” subscale which reflects the high internaize stigma. This results are in concordance with findings from a study conducted in Morocco where 51.9% of their study population reported high score of internal stigma. Furthermore, internal stigma impacted many aspects of life, as personal and social relationships interms of not getting married, social exclusion), career-related (not applying for jobs/training, leave jobs/training) and health-related (avoiding to request care when needed) [27].

Furthermore, most of the enrolled PLWH participants had the feeling that they are not good as others because of their HIV status as they always feel shame. This feeling ranged between *“sometimes”* to *“always felt not good as others”*. This result is similar to findings from a multicenter research conducted in 8 centers in Egypt by “The Egyptian Society for Population Studies and Reproductive Health” (ESPSRH). In this study, over 529 (75.4%) PLWH participants felt shamed and as not good as others because of the illness. Moreover, the internal stigma impacted the quality of life of more than 50% of the participants, who decided to isolate themselves from their family and friends and not to attend social gatherings [28].

According to the present study, men had a higher score in “*Negative self-image”* subscale than women. This was similar to a study conducted in Ethiopia on 412 PLWH where the stigma score was higher among male participants compared to females [29].

PLWH experience regular discrimination. In a study conducted in a number of Arab countries, more than 60% of women in Algeria and Sudan, and more than 70% in Egypt, Jordan, and Yemen, admitted that they would not buy vegetables from a shopkeeper who is living with HIV. Additionally, more than 50% of the surveyed people in the same study who are living with HIV in Algeria reported being denied health services due to their HIV status [30].

The present study has a number of limitations. First, we were unable to assess stigma in multicentrs providing HIV care. however, HIV unit in Alexandria fever hospital is a pooling unit serving all cities in northern Egypt. Furthermore, because we conducted a onetime point cross-sectional study, we were unable to evaluate changes in stigma and discrimination among the study participants over different stages of HIV management.

Despite the limitations, this study has many strengths. To the best of our knowledge, this is the first study with a substantial sample size to address stigma among PLWH in Egypt. This study adds important knowledge about stigma among PLWH, in conjunction with other studies conducted in regional countries which also measured stigma level.

## Conclusion

These results further confirm the fact that PLWH in Egypt experience significant degree of stigma. These data can guide future health interventions to provide psychological and social support for PLWH in our community. Hopefully, this will enhance adherence and, in turn, health outcomes for PLWH.

## Data Availability

All data produced in the present study are available upon reasonable request to the authors

## Acknowledgements

We would like to acknowledge the study participants for accepting to participate in the study.

## Disclosure statement

### Ethical considerations

The study was approved by the institutional review board and the Ethics Committee of the Medical Research Institute, Alexandria University [ref. no.18-2022/21]. All procedures performed in this study involving human participants were in accordance with the ethical standards of the institutional and/or national research committee and with 1964 Helsinki Declaration and its later amendments or comparable ethical standards. Informed written consent was obtained from each participant after explaining the aim and concerns of the study/ Data sheets were coded with numbers to maintain the anonymity and confidentiality of patient’s data.

This article does not contain any studies with animals performed by any of the authors.

## Informed consent

Declared

## Data availability

All data are fully available without restriction by the corresponding author.

## Consent for publication

All authors have read and approved the final manuscript.

## References

1. Goffman E (1963) Stigma: notes on the Management of Spoiled Identity (kindle edition). Touchstone.

2. Link BG, Phelan JC (2001) Conceptualizing stigma. Annual review of Sociology 27: 363–385.

3. Turan B, Budhwani H, Fazeli PL, Browning WR, Raper JL, et al. (2017) How does stigma affect people living with HIV? The mediating roles of internalized and anticipated HIV stigma in the effects of perceived community stigma on health and psychosocial outcomes. AIDS and Behavior 21: 283–291.

4. Li J, Mo PK, Wu AM, Lau JT (2017) Roles of self-stigma, social support, and positive and negative affects as determinants of depressive symptoms among HIV infected men who have sex with men in China. AIDS and Behavior 21: 261–273.

5. Earnshaw VA, Chaudoir SR (2009) From conceptualizing to measuring HIV stigma: a review of HIV stigma mechanism measures. AIDS and Behavior 13: 1160–1177.

6. Deacon H (2005) Understanding HIV/AIDS stigma: A theoretical and methodological analysis: HSRC press.

7. Kalichman SC, Katner, Moira O (2020) HIV-related stigma and non-adherence to antiretroviral medications among people living with HIV in a rural setting. Social Science & Medicine 258: 113092.

8. Mukumbang FC, Mwale JC, van Wyk B (2017) Conceptualising the Factors Affecting Retention in Care of Patients on Antiretroviral Treatment in Kabwe District, Zambia, Using the Ecological Framework. AIDS Research and Treatment 2017: 1–11.

9. Gökengin D, Doroudi F, Tohme J, Collins B, Madani N (2016) HIV/AIDS: trends in the Middle East and North Africa region. International Journal of Infectious Diseases 44: 66–73.

10. Karamouzian M, Madani N, Doroudi F, Haghdoost AA (2017) Improving the quality and quantity of HIV data in the Middle East and North Africa: key challenges and ways forward. International journal of health policy and management 6: 65.

11. UNAIDS (2020) Global AIDS Monitoring 2020: Country progress report - Egypt

12. Ahram (2022) 22,000 people living with AIDS in Egypt: Health ministry alhram.

13. WHO (2022) Global situation and trends.

14. Global AIDS Monitoring (2019) Country progress report -Egypt.

15. Ministry of Health and Population [Egypt], El-Zanaty and Associates [Egypt], and ICF International (2015) Egypt Demographic and Health Survey 2014. Cairo, Egypt and Rockville, Maryland, USA: Ministry of Health and Population and ICF International.

16. Hintze J (2013) NCSS 9. NCSS, LLC. Kaysville, Utah, USA.

17. Berger BE, Ferrans CE, Lashley FR (2001) Measuring stigma in people with HIV: Psychometric assessment of the HIV stigma scale¶. Research in nursing & health 24: 518–529.

18. Reinius M, Wettergren L, Wiklander M, Svedhem V, Ekström AM, et al. (2017) Development of a 12-item short version of the HIV stigma scale. Health and Quality of Life Outcomes 15: 1–9.

19. Younan L, Clinton M, Fares S, Samaha H (2019) WHO Guidelines on Translation and cultural adaptation validity of the Actual Scope of Practice Questionnaire. East Mediterr Health J 25: 181–188.

20. Caiola C, Docherty SL (2018) Black mothers living with HIV picture the social determinants of health. Journal of the Association of Nurses in AIDS Care 29: 204–219.

21. Effah ES (2018) Factors Affecting Adherence to Antiretroviral Therapy Among HIV/AIDS Patients Receiving Treatment at Selected Poly Clinics in Accra Metropolis: University of Ghana.

22. Elopre L, Hook EW, Westfall AO, Zinski A, Mugavero MJ, et al. (2015) The role of early HIV status disclosure in retention in HIV care. AIDS Patient Care and STDs 29: 646–650.

23. Angelo AT, Alemayehu DS (2021) Adherence and its associated factors among adult HIV-infected patients on antiretroviral therapy in South Western Ethiopia, 2020. Patient preference and adherence 15: 299.

24. Dessie G, Wagnew F, Mulugeta H, Amare D, Jara D, et al. (2019) The effect of disclosure on adherence to antiretroviral therapy among adults living with HIV in Ethiopia: a systematic review and meta-analysis. BMC infectious diseases 19: 1–8.

25. Izudi J, Kadengye F (2021) Effect of disclosure of HIV status on patient representation and adherence to clinic visits in eastern Uganda: A propensity-score matched analysis. Plos one 16: e0258745.

26. Tshweneagae VM, Mgutshini T (2015) Disclosure of HIV status to sexual partners by people living with HIV. curationis 38: 1–6.

27. Moussa AB, Delabre RM, Villes V, Elkhammas M, Bennani A, et al. (2021) Determinants and effects or consequences of internal HIV-related stigma among people living with HIV in Morocco. BMC Public Health 21: 1–11.

28. Khattab HA, El-Geneidy M, Gamil F, Gaballah M (2013) Stigma experienced by people living with HIV in Egypt. The Egyptian Society for Population Studies and Reproductive Health (ESPSRH) 40 p.

29. Ataro Z, Abrham T (2020) Gender differences in perceived stigma and coping strategies among people living with HIV/AIDS at jugal hospital, Harar, Ethiopia. Psychology Research and Behavior Management: 1191–1200.

30. UNAIDS (2018) Global aids Updates.

