## Supplementary material for "Stigma among people living with HIV in Northern Egypt": tables

**List of tables**

**Table 1: Demographic and HIV related characteristics of the enrolled PLWH**

| Characteristics | n (%) |
| --- | --- |
| Age |  |
| 19 – <30 Years | 161 (20.5) |
| 30 – <50 | 532 (67.8) |
| ≥ 50 | 92 (11.7) |
| Sex |  |
| Male | 567 (72.2) |
| Female | 218 (27.8) |
| Marital status |  |
| Married | 309 (39.4) |
| Single | 278 (34.4) |
| Divorced | 100 (12.7) |
| Widow | 98 (12.5) |
| Residence |  |
| Alexandria | 668 (85.1) |
| Other cities^a^ | 117 (14.9) |
| Education |  |
| Low literacy^b^ | 90 (11.5) |
| High literacy^c^ | 695 (88.5) |
| Employment |  |
| Student or employee | 410 (52.2) |
| Housewife or retired | 200 (25.5) |
| Owner | 105 (13.4) |
| Unemployed | 70 (8.9) |
| HIV related risky behavior |  |
| Heterosexual, multiple partners | 417 (53.1) |
| Unknown risk /other risks | 114 (14.5) |
| Intravenous Drug user | 106 (13.5) |
| MSM | 100 (12.7) |
| Blood transfusion | 48 (6.2) |

a; nearby cities in northern Egypt

b; Illiteracy, read and write

c; 6 years of education or higher

PLWH; people living with HIV

MSM; men who have sex with men

| Item | n (%) | Median item  Score (IQR)^a^ | Reliability  α |
| --- | --- | --- | --- |
| 1- Personal stigma  P1- Some people avoid touching me once they know I have HIV  P2- People I care about stop calling after knowing that I have HIV  P3- I lost my friends because I told them that I have HIV | 415 (52.9)  415 (52.9)  415 (52.9)  414 (52.7) | 1 (1-2)  1 (1-1)  1 (1-1) | 0.941 |
| 2- Disclosure concerns  D1- Telling anyone that I have HIV is a risk  D2- I work hard to keep my HIV a secret  D3- I am very careful about telling anyone I have HIV (work) | 785 (100.0)  783 (99.7)  785 (100.0)  562 (71.6) | 4 (4-4)  4 (4-4)  4 (4-4) | 0.882 |
| 3- Concerns about public attitudes  C1- People with HIV are treated like outcasts  C2- Most people believe that the person who has HIV is a bad behaved person  C3- Most people do not feel comfortable around someone has HIV | 784 (99.9)  777 (99.0)  784 (99.9)  777 (99.0) | 4 (4-4)  4 (4-4)  4 (4-4) | 0.870 |
| 4- Negative self-image  N1- I feel guilty because I have HIV  N2- People's attitudes toward HIV make me feel worse about myself  N3- I feel that I am not as good as others because I have HIV  Overall | 785 (100.0)  785 (100.0)  764 (97.3)  785 (100.0) | 3 (1-4)  4 (3-4)  3 (3-4) | 0.657  **0.743** |

**Table 2: Description of the 12- short version of the HIV stigma scale among the enrolled PLWH**

a; Inter Quartile Range

PLWH; people living with HIV

**Table 3: Association between sociodemographic characteristics and the total stigma score**

| Characteristics | Stigma SCORE Median (min-max) | *p* value¬ |
| --- | --- | --- |
| Sex |  | <0.001 ^a^ |
| Male | 34 (12-48) |  |
| Female | 32 (21-47) |  |
| Residence |  | 0.033 ^a^ |
| Alexandria | 33 (21-48) |  |
| Other cities^c^ | 32 (12-47) |  |
| Education |  | 0.282 ^b^ |
| Illiteracy, read and write | 32.5 (12-48) |  |
| ≤ 12 years of education | 34 (21-48) |  |
| University education or higher | 33 (22-47) |  |

^a^: by Mann-Whitney U test

^b^: by Kruskal-Wallis test

c; nearby cities in northern Egypt

¬*p* <0.1 is set as a level of significance

Figure legend

Figure 1: Median score of the different stigma subscales

Figure 2: Distribution of the median score of stigma subscales by sex

Figure 3: Distribution of median stigma score of the four subscales by age category

Figure 4: Questions with the highest score of “4” points in each subscale
